# Plasma Cell-Free RNA Captures Immune Dynamics and Predicts GVHD after Hematopoietic Stem Cell Transplantation

**DOI:** 10.1101/2024.05.15.24307448

**Authors:** Conor Loy, Glory Agun, Matthew P. Cheng, Isabel H. Gonzalez-Bocco, Joan Lenz, Emma Belcher, Andrew Bliss, Daniel Eweis-LaBolle, Tinyi Chu, Joseph Pidala, Jerome Ritz, Iwijn De Vlaminck

## Abstract

Despite long-standing success of hematopoietic stem cell transplantation (HSCT) in the treatment of blood cancers and severe immune disorders, monitoring post-transplant complications remains challenging, in part because of the limited availability of informative molecular tests. Here, we evaluated the utility of cell-free RNA (cfRNA) in plasma as an analyte to monitor HSCT recipients and predict immune-related complications. We used RNA-sequencing to profile cfRNA in 549 plasma samples collected at predetermined time points from allogeneic HSCT recipients at two transplant centers. We found that cfRNA temporal profiles in recipients with a complication-free course follow a relatively uniform trajectory, whereas recipients who experience complications exhibit divergent trajectories. cfRNA patterns also differed greatly from circulating cell counts, providing complementary information on cell expansion and turnover. We also used cfRNA profiles at early time points to train machine learning models for the early prediction of acute GVHD, chronic and late acute GVHD, and the outcome of immunosuppression taper. We discovered shared signatures that achieved good performance across different scenarios in both cohorts. Overall, our findings provide support for the use of plasma cfRNA profiling to monitor immune related complications of HSCT.

## Main Text

Hematopoietic stem cell transplantation (HSCT) is one of the oldest and most widely used immunotherapies (*1*). In allogeneic HSCT, the recipient’s hematopoietic and immune systems are rapidly replaced by donor cells. For recipients, the engraftment of donor immune cells contributes to a graft-versus-tumor effect (*2*, *3*). However, the success of HSCT is limited by the frequent occurrence of complications, including graft-versus-host disease (GVHD), relapse, and infection (*4*–*6*). Most HSCT recipients experience at least one complication, and complications can often co-occur (*7*). Complications can manifest with non-specific symptoms, such as rash, elevated liver enzymes, or diarrhea, and progress rapidly, making early prediction and diagnosis critical. GVHD is one of the most common and severe complications of allogeneic HSCT, arising when engrafted T cells attack the host’s organs (*6*). This can lead to tissue damage and in severe cases, death. GVHD is categorized into acute GVHD (aGVHD), which typically occurs within the initial 100 days post-transplantation, and chronic GVHD (cGVHD), which often develops later and can persist for years (*8*–*10*). The current standard of care involves a battery of diagnostic tests including invasive tissue biopsies (*11*, *12*). There is a clear need for noninvasive molecular tests to monitor HSCT recipients and predict complications before they occur (*13*–*15*).

Cell-free DNA (cfDNA) and cell-free RNA (cfRNA) are released in plasma via active excretion and as a byproduct of cell death, and thus reflect cell turnover, immune activity, and cellular damage (*16*). While previous studies have shown that methylation profiling of cfDNA can identify HSCT-related complications (*17*–*19*), and that cfRNA profiles immediately after HSCT reflect hematopoietic recolonization (*20*), the long-term dynamics of cfRNA following HSCT and its potential to predict complications remain unexplored. In this study, we implemented plasma cfRNA profiling by RNA-sequencing on 549 samples across two cohorts to explore HSCT dynamics and prognostic signatures of GVHD. Cohort 1 includes 449 samples collected at predefined time points before and after HSCT, while cohort 2 consists of 100 samples from recipients undergoing immunosuppression taper. As a control group, we also analyzed cfRNA from 18 healthy stem cell donors. We found that recipients with complication-free recoveries followed a shared path toward health, while recipients who experienced complications exhibited more molecularly heterogeneous recoveries. We also found that cfRNA profiles at engraftment reflect future aGVHD and discovered shared signatures between cohort 1 and cohort 2 that are prognostic of GVHD. This study leverages innovative methodology and machine learning to discover signatures of GVHD across cohorts, warranting further exploration and clinical validation of cfRNA in HSCT.

## Results

### cfRNA dynamics associated with HSCT

To assess the utility of cfRNA to monitor HSCT treatment and recovery, we prospectively collected and analyzed 449 plasma samples from 96 subjects who underwent allogeneic HSCT at the Dana-Farber Cancer Institute (DFCI) for a variety of conditions, including leukemia, lymphoma, and aplastic anemia (**Table S1**). Samples were collected longitudinally at predetermined time points before and after HSCT: prior to conditioning therapy, on the day of and prior to infusion (day 0), at hematopoietic engraftment, and at one, two, three and six months after transplantation (**Fig. 1A** and **B**). We also collected and analyzed plasma samples from 18 healthy stem cell donors. Complications were frequent and included acute and chronic GVHD (aGVHD 45%, cGVHD 53%), BK virus reactivation (BKVR, 45%), cancer relapse (40%), and graft failure (6%). Many recipients experienced more than one complication, and the timing of these events differed both between individuals and among complication categories (**Fig. S1A-B**).

**Fig. 1.**
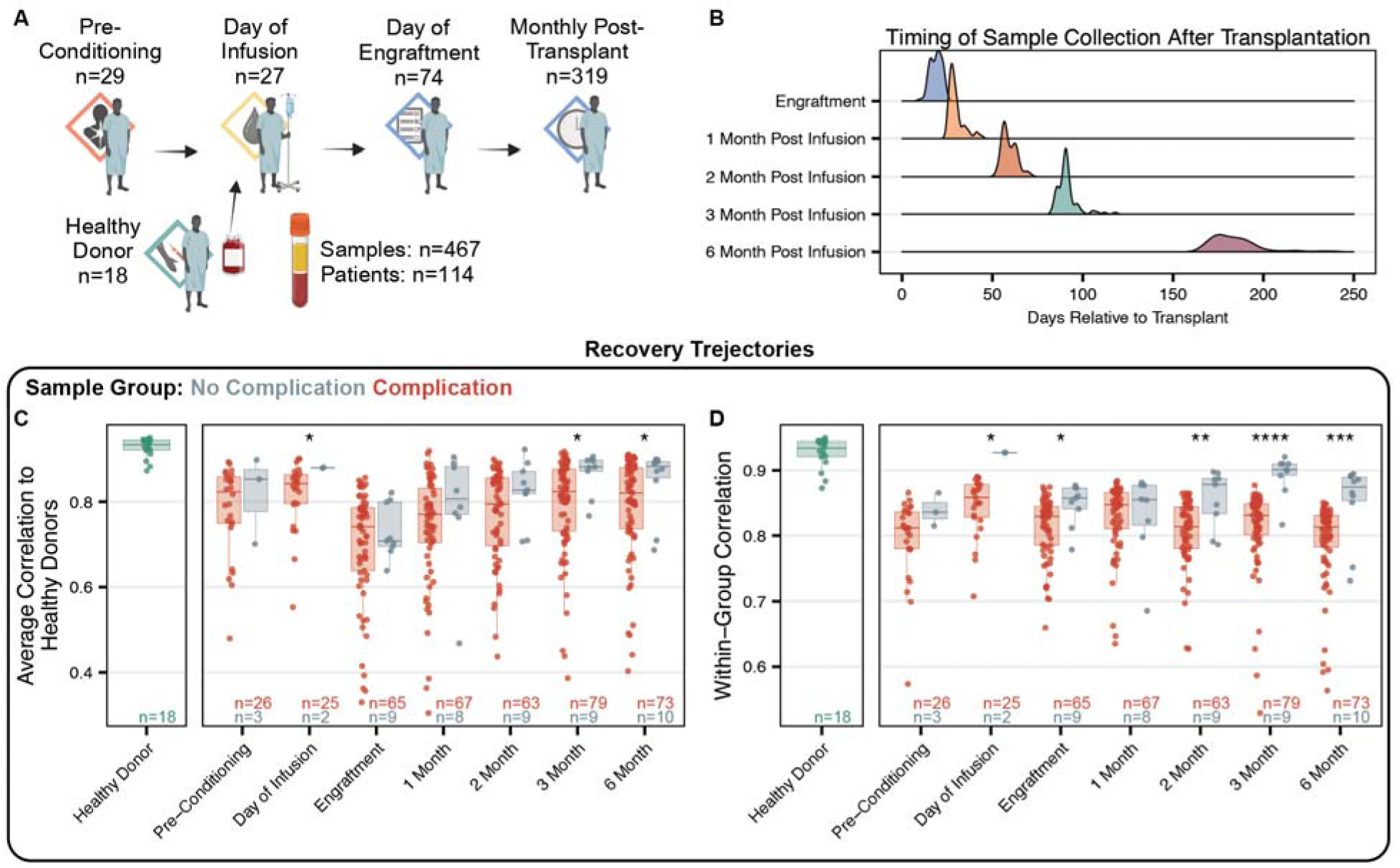
Sample cohort overview. (**A**) Sample collection overview. (**B**) Day relative to transplantation samples were collected for each timepoint. (**C**) Correlation of cfRNA abundance to healthy stem cell donors of samples from with and without complications (**Methods**). (**D**) Within group average correlations of recipients who did and did not experience complications across the sampling time course (Pearson correlation). Self-comparisons were removed prior to average calculation. Asterisks indicate statistical significance by Mann-Whitney U test using p-values as follows: ns, non-significant; *, p < 0.1; **, p < 0.01; ***, p < 0.001; ****, p < 0.001.

### cfRNA reflects recovery and return to health

To determine if cfRNA can measure a “distance to normal”, we quantified the average correlation of the patient profiles to the healthy donor reference profiles (**Fig. 1C**, **Methods**). We then divided the recipients into two broad categories: those who experienced ≥1 major complication within the first nine months post-transplant (n=82) and those who did not (n=10, **Methods**). We found that cfRNA profiles at the engraftment time point showed the lowest similarity to normal profiles, followed by a time-dependent increase in correlation toward the normal reference. Notably, recipients with complications exhibited lower correlation than those without complications, with significant differences at the three- and six-month time points (p-value < 0.1). We observed similar trends when quantifying the distance to normal at the level of cell-types-of-origin (**Fig. S1C**). We also measured within-group correlations at each time point and found that recipients with complications displayed significantly less internal consistency compared to both complication-free recipients and healthy donors starting at the two-month time point (p-value < 0.1, **Fig. 1D**). This pattern indicates that recipients with a complication-free course follow a relatively uniform trajectory, whereas recipients who suffer complications each exhibit a unique trajectory, an observation reminiscent of the Anna Karenina principle in statistics (*21*).

### cfRNA origins are dynamic and distinct from whole blood cell counts

To gain further understanding of the origins of plasma cfRNA profiles during HSCT, we quantified the cell types that contribute cfRNA to the plasma mixture. For this purpose, we used BayesPrism (*22*) to computationally deconvolve cfRNA based on the *Tabula Sapiens* single-cell atlas (*23*) as a reference (**Fig. 2A**). Across all recipients, regardless of complication status, this analysis revealed rich dynamic changes in the cell-types-of-origin of cfRNA in response to conditioning and stem cell infusion (**Fig. 2B**). For example, the proportion of hematopoietic-lineage-cell derived cfRNA was much lower on the day of infusion than at engraftment, likely reflecting the impact of pre-transplant conditioning and the subsequent recovery of donor hematopoiesis. Interestingly, cell-type-of-origin patterns differed between patients who were HLA haploidentical to their donors and those who were not, consistent with differences in conditioning and immunosuppression regimens.

**Fig. 2.**
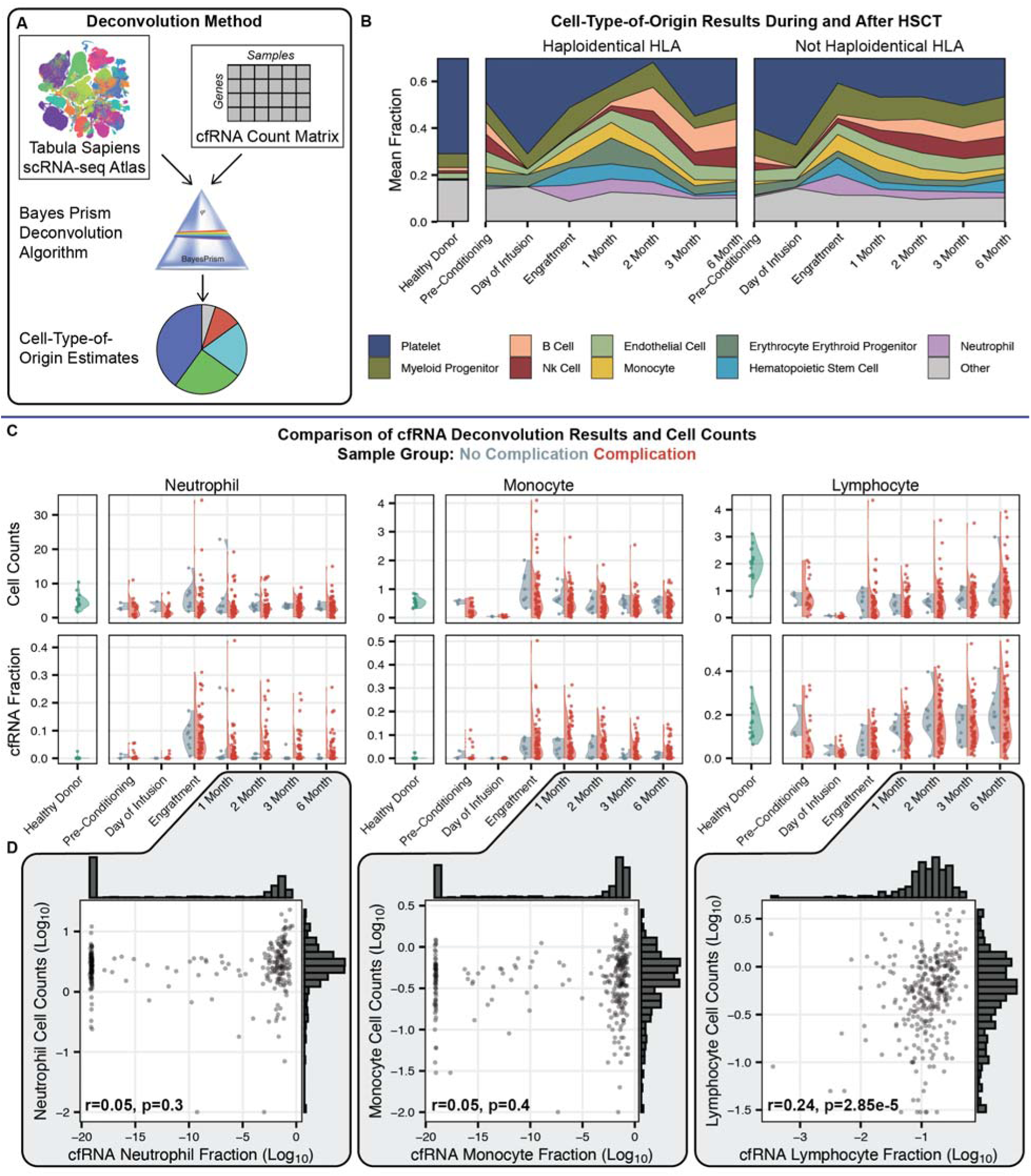
cfRNA origins during and after HSCT. (**A**) Overview of deconvolution method. (**B**) Stacked area plot of mean fraction of top ten most abundant cell types across the time course, split by HLA matching between donor and patient. (**C**) Complete blood cell counts and cfRNA cell type of origin fraction for paired samples across the treatment time course. Samples are colored by grouping: those that did not experience a complication in the first 9 months and those that did. (**D**) Scatterplots of cfRNA cell-type-of-origin estimates and complete blood cell counts with accompanying histograms for each axis for neutrophils, monocytes, and lymphocytes. Pearson correlation was calculated for each cell type and the resulting statistics are shown. Plot created and correlation calculated after removal of a single outlier in the lymphocyte analysis.

When we compared cfRNA cell-type-of-origins with blood cell counts obtained via automated image-based cell counting key differences emerged (paired samples, **Fig. 2C-D**, **Methods**). While lymphocyte cfRNA fractions tracked lymphocyte cell counts, this relationship did not hold true for neutrophils or monocytes (**Fig. 2C**). Specifically, recipients who developed complications exhibited marked increases in neutrophil- and monocyte-derived cfRNA without a corresponding rise in cell counts. After engraftment, log-normalized neutrophil and monocyte cell counts followed a unimodal distribution, whereas log-normalized cfRNA-derived neutrophil and monocyte fractions were bimodal (**Fig. 2D**). We attribute these differences to the fact that cfRNA profiles reflect both cell-type abundance and turnover rate. The lack of correlation between cfRNA levels and whole-blood counts for neutrophils and monocytes suggests an increased turnover rate in patients with complications.

### cfRNA signatures of GVHD

Acute GVHD (aGVHD) is an early onset complication of HSCT in which engrafted T cells attack the recipient’s tissues. aGVHD frequently occurs within the first 100 days post-transplantation (**Fig. 3A**). Given this early onset, we investigated cfRNA profiles at engraftment for predictive signatures of aGVHD, excluding samples from one recipient who developed aGVHD before engraftment (**Methods**). We observed that cfRNA profiles at engraftment in recipients who later developed aGVHD differed from those in recipients who did not in several keyways. First, these profiles were significantly less similar to those of healthy donors (**Fig. 3B**; p-value = 0.049). They also showed higher contributions from leukocytes (**Fig. 3B**; p-value = 0.03), greater overall diversity in cell-type origins (Simpson diversity, p-value = 0.016), and higher levels of histone transcripts (**Fig. 3B**; p-value = 0.06). Last, at the gene level, we identified a total of 296 differentially abundant transcripts (**Data S1**, DESeq2, Benjamini-Hochberg (BH) adjusted p-value < 0.1). Pathway analysis (Qiagen IPA) revealed that genes elevated in recipients who developed aGVHD were enriched in pathways related to cell expansion and immune activation (**Fig. S2A**). When we clustered the samples using the most differentially abundant transcripts (BH adjusted p-value < 0.1, area under the curve of the receiver operating characteristic (AUC-ROC) > 0.65), samples separated by future aGVHD status (**Fig. 3C**). Finally, we investigated transcript abundance relative to the time to aGVHD diagnosis using samples collected at engraftment, one month, and two months before diagnosis. Doing so, we identified 119 transcripts significantly associated with time to diagnosis (**Data S2**, **Methods**; DESeq2, BH adjusted p-value < 0.1). Notably, we found that the abundance of Major Histocompatibility Complex transcripts, Class I - A, B, C, and E (*HLA-A*, *HLA-B, HLA-C*, and *HLA-E*) were all negatively correlated with time to aGVHD diagnosis: the closer the sample was taken to aGVHD diagnosis, the higher the abundance of these HLA transcripts (**Fig. S2B**). This trend may reflect an escalating immune response in the early stages of aGVHD.

**Fig. 3.**
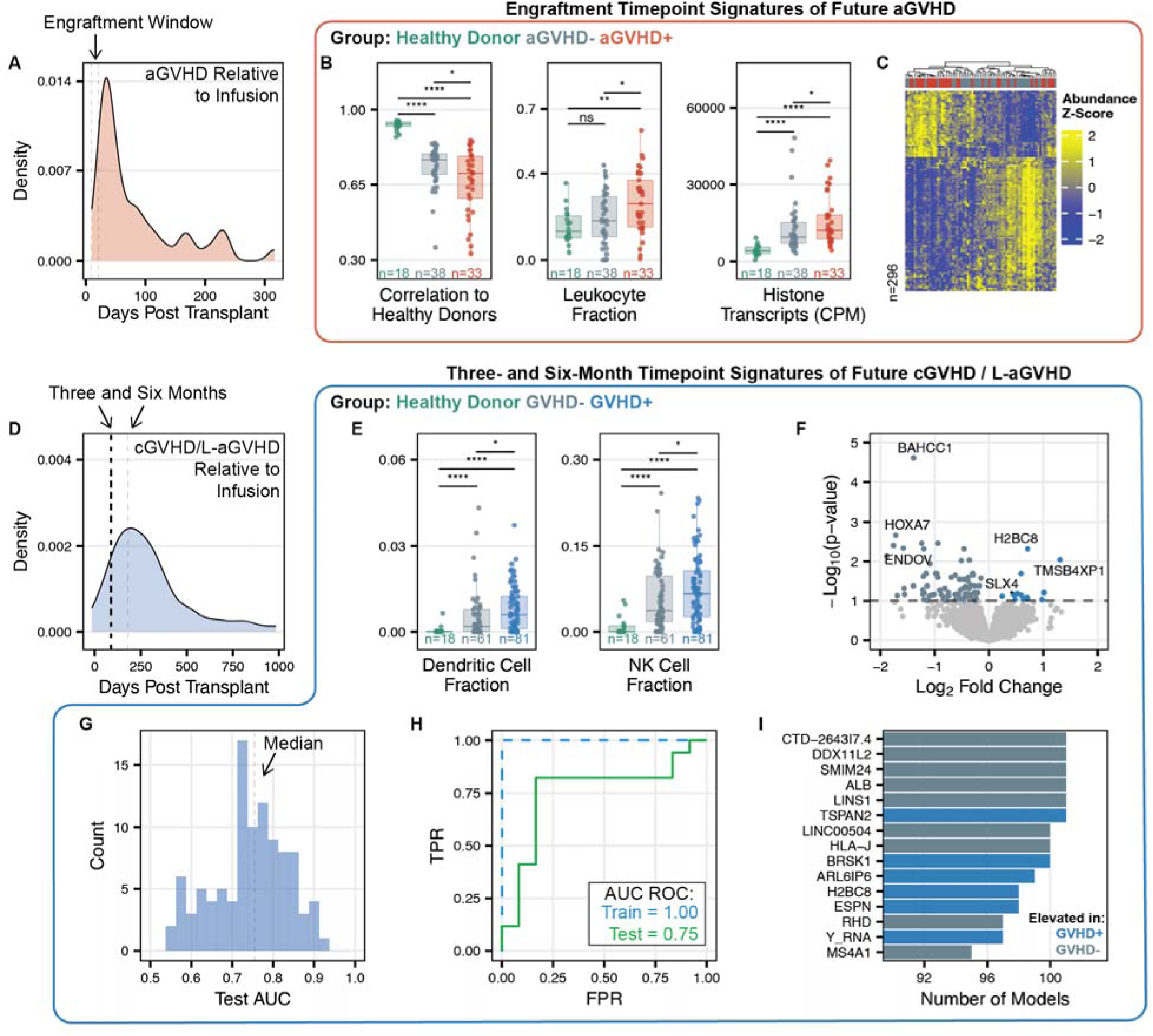
cfRNA signatures prior to GVHD diagnosis. (**A**) Number of days post infusion of aGVHD diagnosis. (**B**) Correlation of cfRNA abundance to healthy stem cell donors, leukocyte fraction, and sum of replication histone transcripts of samples from recipients who develop aGVHD and those who do not. Samples from prior to aGVHD diagnosis. (**C**) Scaled variance stabilization transformation values of significantly differentially abundant genes at the engraftment time point of samples from recipients that develop aGVHD and those that do not develop (DESeq2, BH-adjusted p-value < 0.1, abundance AUC-ROC > 0.65). Samples and genes clustered based on correlation. (**D**) Number of days post infusion of cGVHD and/or L-aGVHD diagnosis. (**E**) Dendritic cell and NK cell fraction of samples from recipients who develop cGVHD and/or L-aGVHD and those who do not. Samples from prior to cGVHD and/or L-aGVHD diagnosis. Asterisks indicate statistical significance: ns, non-significant; *, p < 0.1; **, p < 0.01; ***, p < 0.001; ****, p < 0.001. (**F**) Volcano plot of the results from the differential abundance analysis comparing engraftment time point samples from recipients who develop cGVHD and/or L-aGVHD and those who do not (DESeq2, BH-adjusted p-values). (**G**) Distribution of test set AUC-ROC scores across all 101 repeated train-test splits. (**H**) AUC-ROC curves of the training and test set of the median performing model. (**I**) The top 15 most selected transcripts of the 101 repeated stratified train-test split modeling. Asterisks indicate statistical significance: ns, non-significant; *, p < 0.1; **, p < 0.01; ***, p < 0.001; ****, p < 0.001.

Chronic GVHD (cGVHD) and late acute GVHD (aGVHD) are later-onset complications of HSCT (**Fig. 3D**). To identify cfRNA signatures predictive of these complications, we analyzed samples collected at the three- and six-month time points prior to cGVHD or late aGVHD diagnosis (**Methods**). We again found significant differences for patients who developed GVHD. First, recipients who developed GVHD exhibited significantly higher levels of dendritic cell and NK cell-derived cfRNA compared to those who did not develop GVHD (**Fig. 3E**; p-value < 0.1). At the transcript level, we identified 106 significantly differentially abundant transcripts (**Fig. 3F**, **Data S3,** BH-adjusted p-value < 0.1). Given the large number of samples available (n=142), we explored the potential to train a machine learning model to predict GVHD. We performed 101 repeated stratified train-test splits without replacement to train and test LASSO regression models (**Methods**). The model with the median test performance achieved good performance (AUC-ROC train = 1.00, test = 0.75; **Fig. 3G-H**). Notably, a small subset of transcripts was repeatedly selected across iterations by the LASSO algorithm, with six transcripts (*CTD-264317.4, DDX11L2, SMIM24, ALB, LINS1, TSPAN1*) represented in all 101 models **(Fig. 3I**). Interestingly, multiple genes used for blood typing and matching, *RHD, HLA-J,* and *ABO*, were selected by many models.

### cfRNA signatures for risk of immunosuppression taper failure

Immunosuppression is discontinued for many allogeneic HSCT recipients to achieve immune tolerance (*24*, *25*). Predicting which recipients can successfully undergo immunosuppression tapering without developing GVHD is a major clinical challenge, as there are no established predictive biomarkers. To assess the utility of cfRNA to predict the outcome of immunosuppression taper, we collected and analyzed 100 samples from 53 subjects who underwent allogeneic HSCT at the Moffitt Cancer Center (MCC) for a range of conditions (**Fig. 4A-B, Table S2**). We obtained samples at the following key time points: the first day of immunosuppression taper, the day of immunosuppression stop, the day of taper failure (GVHD, during or after taper), and at two weeks, one month, three months, and six months after the end of taper (**Fig. 4A**). The initiation of taper varied among recipients based on individual clinical decision making, and the onset of taper failure occurred across a broad range of days following the start of taper (**Fig. 4B**). Based on these outcomes, recipients were classified as either having failed taper (n = 30, GVHD occurring during taper or after immunosuppression stop) or successfully completed taper (n = 23, no GVHD during/after taper and remained GVHD free).

**Fig. 4.**
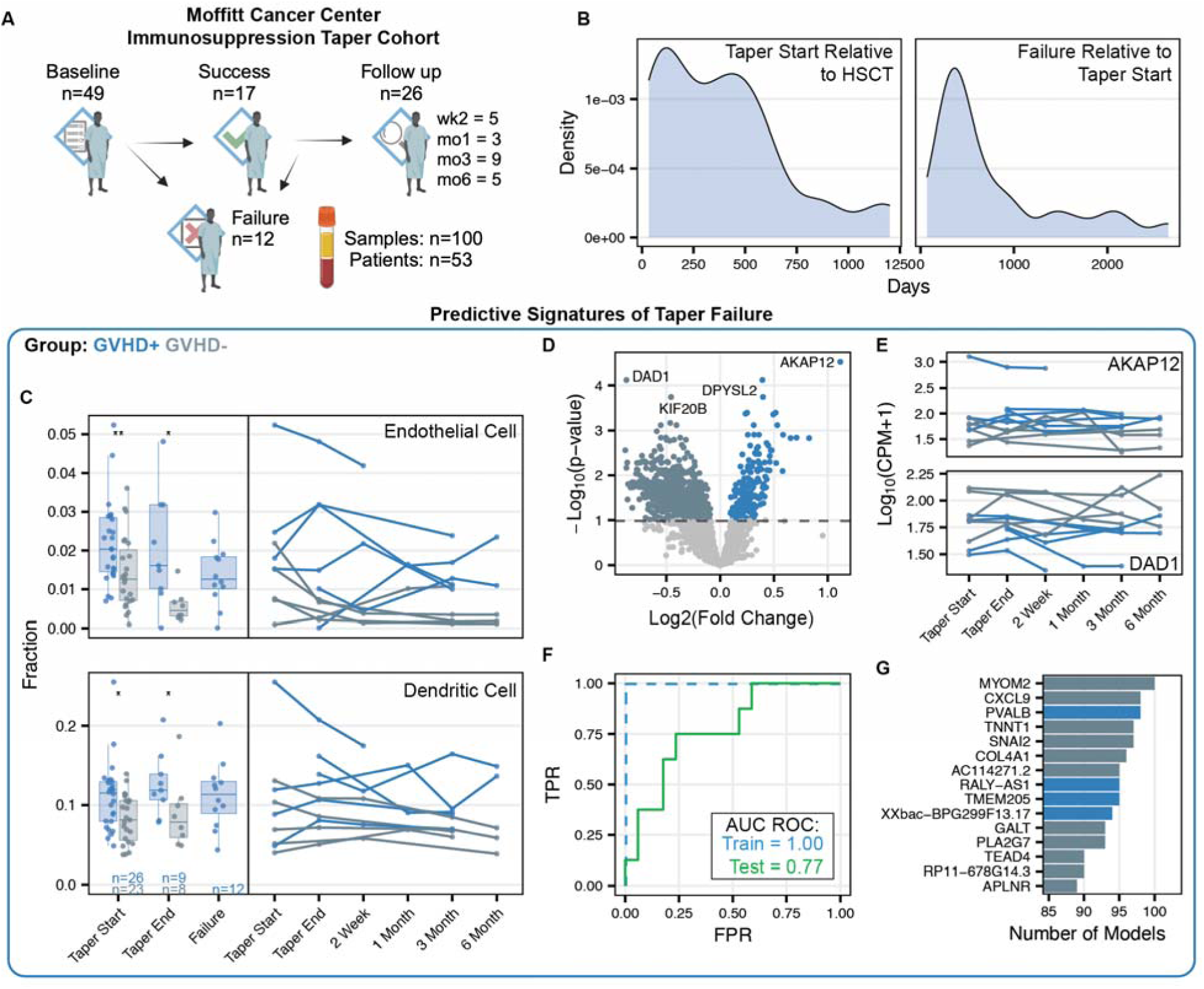
Immunosuppression Taper. (**A**) Clinical cohort overview. (**B**) Distribution of days after HSCT that immunosuppression taper started and days after taper start that failure occurred. (**C**) cfRNA cell-type-of-origin measurements of mature conventional dendritic cell and endothelial cell at the start, end, and failure of taper, along with temporal dynamics in recipients with more than one sample. Asterisks indicate statistical significance by using p-values as follows: ns, non-significant; *, p < 0.1; **, p < 0.01; ***, p < 0.001; ****, p < 0.001. (**D**) Volcano plot of the results from the differential abundance analysis comparing all samples from recipients who failed immunosuppression taper and those that did not (DESeq2, BH-adjusted p-values). (**E**) Longitudinal CPM values of *AKAP12* and *DAD1* in recipients who fail immunosuppression taper and those that do not. (**F**) AUC-ROC curves of the training and test set of the median performing model. (**G**) The top 15 most selected transcripts of the 101 repeated stratified train-test split modeling. Asterisks indicate statistical significance: ns, non-significant; *, p < 0.1; **, p < 0.01; ***, p < 0.001; ****, p < 0.001.

We first aimed to identify cfRNA signatures predictive of immunosuppression taper failure at the start of taper, given the potential impact of such signatures on clinical decision-making. Recipients who later failed taper had higher levels of dendritic cell– and endothelial cell–derived cfRNA at the onset of taper (**Fig. 4C**; p-value < 0.1), a pattern also observed at the end of taper and at the time of failure in event-driven collections. Longitudinal analysis further revealed that these elevated cfRNA signals remained consistently high post-taper in recipients who ultimately develop GVHD. To identify transcript-level differences, we expanded our comparison by including all available pre-failure samples. We found 836 differentially abundant transcripts (**Fig. 4D, Data S4**; DESeq2, BH-adjusted p-value < 0.1). Of these, the two most significantly differentially abundant were *DAD1* and *AKAP12*, which relate to cell death regulation and cell adhesion (**Fig. 4E**). We next trained a machine learning model (n=88) to predict immunosuppression failure (LASSO regression model, 101 repeated stratified train-test splits without replacement, **Methods**). Considering the longitudinal design of the study, we used stratified group splitting for the train/test partitioning. This approach ensured that all samples from an individual recipient were assigned exclusively to either the training or test set, thereby avoiding data leakage. The model with the median test performance achieved promising results (AUC-ROC train = 1.00, test = 0.77; **Fig. 4F**). Finally, we asked which transcripts were most consistently chosen by the LASSO algorithm. A subset of transcripts was repeatedly selected across iterations, with three transcripts (*MYOM2, CXCL9, PVALB*) represented in over 96 models **(Fig. 4G**).

### Shared signatures of GVHD

To identify shared cfRNA signatures of GVHD across the DFCI and MCC cohorts, we analyzed overlap in transcripts selected by the repeated stratified machine learning analyses. Many features overlapped, despite differences in collection timing and sample handling, with *RHD* and *CCL21* being the most frequently selecting (**Fig. 5A**). In both cohorts *RHD* was more abundant in recipients without GVHD, while *CCL21* was more abundant in recipients who developed GVHD. Based on this, we created a cfRNA score (CCL21/RHD), which showed promising performance across both cohorts (AUC-ROC MCC=0.71, DFCI=0.68, **Fig. 5B-C**). In a Kaplan-Meier analysis, recipients with CCL21/RHD scores greater than the median developed GVHD earlier than those with lower scores lower than the median (**Fig. 5D**). This analysis treated GVHD as the primary event, without accounting for competing risks like death or relapse. Finally, analyzing the expression of these transcripts in the *Tabula Sapiens* scRNA-seq atlas (*23*), we found that *RHD* is primarily expressed in erythroid progenitor cells, while *CCL21* is almost exclusively expressed in endothelial cells of lymphatic vessels (**Fig 5E-F**).

**Fig. 5.**
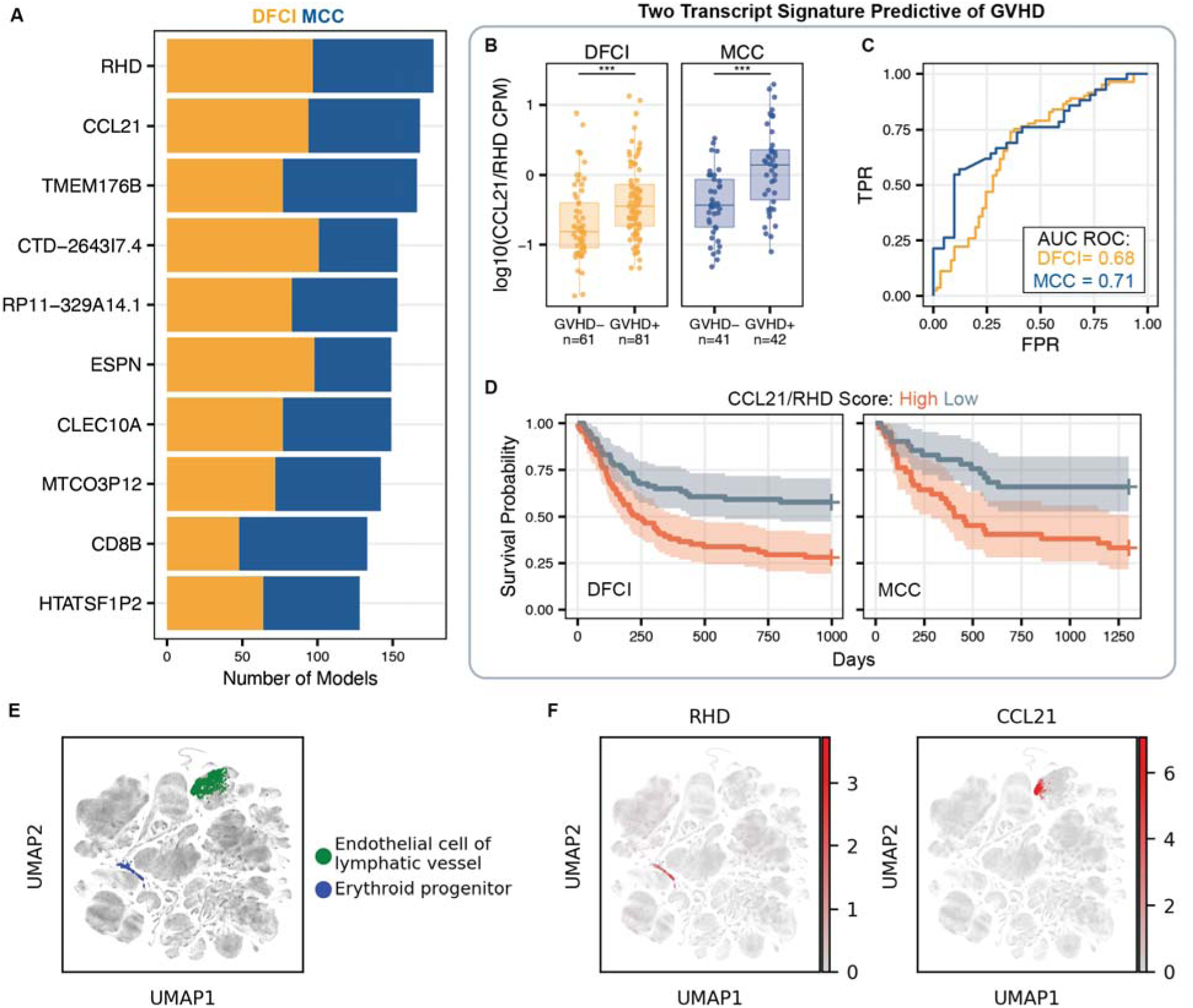
Shared signatures between cohorts. (**A**) The most selected transcripts from the analyses shown in Fig. 4F and Fig. 5G. (**B**) Values of *CCL21*/*RHD* CPM log10 scaled in three- and six-month samples from DFCI and all samples from MCC. Asterisks indicate statistical significance: ns, non-significant; *, p < 0.1; **, p < 0.01; ***, p < 0.001; ****, p < 0.001. (**C)** ROC curves of the *CCL21*/*RHD* CPM values from MCC and DFCI. (**D**) KM survivorship curves of *CCL21*/*RHD* CPM values split into high and low groups based on medians separately calculated for each cohort. (**E**) UMAP visualization of the *Tabula Sapiens* scRNA-seq atlas. Endothelial cells of lymphatic vessels and erythroid progenitor cells colored. (**F**) Expression levels of *RHD* and *CCL21* in the *Tabula Sapiens* scRNA-seq atlas.

## Discussion

Recent work has shown that cfRNA profiling provides insights into immune activity and organ damage in disease specific patterns, offering a new modality to understand and diagnose inflammatory disease (*16*, *26*–*30*). Motivated by these insights, we evaluated the potential of circulating cfRNA to monitor HSCT across two independent cohorts. We observed dynamic changes in response to therapy and identified signatures associated with GVHD risk across timepoints and clinical settings.

According to Aristotle, "it is possible to fail in many ways, while to succeed is possible only in one way". In line with this old perspective, we found that recipients free of complications follow highly similar cfRNA trajectories approaching normal, while recipients who did experience complications follow unique, individualized trajectories. These observations provide a theoretical framework for addressing similarly heterogeneous patient groups. In addition, from a practical standpoint, we propose that a “distance to health” or correlation to a normal/healthy cfRNA profile might be useful as a gauge of the success of HSCT and as a guide for treatment decisions.

Blood cell counts are commonly used to gauge immune status, yet in the post-HSCT setting they have limited prognostic or diagnostic value. This is partly because cell counts fail to measure immune turnover. For example, immune cell death and replacement may offset each other, resulting in stable cell counts despite potential underlying pathology. By contrast, our findings suggest that cfRNA captures the abundance of immune cell types and their rate of turnover or death, potentially offering deeper insight into underlying immunologic activity. We observed elevated cfRNA from neutrophils and monocytes in recipients with complications, despite stable blood cell counts—indicating heightened cell turnover or activation. Additionally, we found that cfRNA from dendritic and NK cells differed significantly at three- and six-months post-transplant between patients who did and did not develop GVHD. This aligns with prior studies which found increased cytotoxic NK cells in patients who develop GVHD (*31*, *32*). Ultimately, a panel of cfRNA biomarkers reflecting neutrophil, monocyte, dendritic, and other immune cell–derived transcripts could offer an actionable snapshot of transplant status.

GVHD occurs when the transplanted immune system recognizes the recipient’s body as foreign, triggering an immune response that can lead to tissue damage and mortality (*10*, *33*). aGVHD typically develops within 100 days post-transplant, making early biomarkers essential for identifying at-risk recipients. At engraftment, recipients who later develop aGVHD exhibited cfRNA profiles more distinct from healthy donors with signatures suggesting aggressive immune reconstitution and activation, independent of conditioning regimen and graft source. At later time points and during immunosuppressive therapy, we observed increased dendritic cell–derived cfRNA in recipients who developed chronic or late-acute GVHD, reinforcing the role of antigen-presenting cells in disease progression. Machine learning models identified *RHD*, *ABO*, and *HLA-J*—genes involved in antigen presentation and blood typing—as predictive of GVHD, with *RHD* emerging as the most shared signature across cohorts. *RHD*, which encodes the RhD antigen, plays a key role in immune recognition. Clinically, Rh incompatibility during pregnancy can result in hemolytic disease of the fetus, while in transfusion medicine, RhD alloimmunization can cause complications (*34*–*38*). Although Rh antigen matching has been investigated in the context of HSCT, prior studies have found no clear association between RhD mismatches and clinical outcomes, including GVHD (*36*–*38*). Interestingly, our findings indicate that lower levels of cfRNA *RHD* transcripts correlate with the future development of GVHD. Another shared signature, *CCL21*, is a chemokine that binds to the cell-surface receptor CCR7, guiding immune cells to secondary lymphoid organs (*39*). This chemokine-driven migration may promote antigen presentation and the T-cell activation processes that are central to GVHD. We examined the expression patterns of *RHD* and *CCL21* in the *Tabula Sapiens* scRNA-seq atlas and found that *RHD* was primarily expressed in erythroid progenitor cells, while *CCL21* was almost exclusively detected in endothelial cells of lymphatic vessels—both cell types not found in circulation. The expression pattern of *CCL21* is particularly compelling given that thrombotic microangiopathy is associated with GVHD and is an important cause of multi-organ failure and non-relapse mortality (*40*). These findings highlight the unique advantages of cfRNA profiling, as it enables the detection of transcripts from cell types not present in circulation—such as bone marrow-resident erythroid progenitors and solid tissue endothelial cells. Additionally, cfRNA sequencing allows for an untargeted, comprehensive measurement of the transcriptome and resources like *Tabula Sapiens* can be leveraged to better interpret these data, unlike circulating protein measurements. These findings provide insight into GVHD pathogenesis and highlight cfRNA as a promising tool for early GVHD risk assessment.

Our study has several important limitations. First, although we analyzed a sizable number of samples (n=549) across two cohorts, the number of patients included is limited (n=149) and the sample size still does not capture the full spectrum of clinical variability in this highly heterogeneous cohort of patients who suffer myriad complications. Second, GVHD was diagnosed by consensus criteria and not every patient had a biopsy, potentially serving as a source of bias. Ultimately, translating cfRNA signatures into routine clinical practice will require additional validation in larger studies, with prospective enrollment and standardized measurement protocols. In summary, this study introduces analysis techniques that may be broadly useful and highlights the promise of cfRNA to monitor HSCT. Beyond HSCT, these insights may inspire broader applications in cellular immunotherapy, positioning cfRNA as a key bioanalyte to measure immune activity and treatment response across diverse clinical contexts.

## Materials and Methods

### Ethics Statement

The study was approved by the Dana-Farber/Harvard Cancer Center’s Office of Human Research Studies and Moffitt Cancer Center IRB. All recipients provided written informed consent.

### Clinical Cohorts

#### Clinical Cohort 1

Adult recipients undergoing allogeneic HSCT at Dana-Farber Cancer Institute were prospectively enrolled on a rolling basis from August 2018 to August 2019. Samples were collected longitudinally at predetermined time points: prior to conditioning therapy (Pre-Conditioning), at the day of infusion prior to stem cell infusion (Day of Infusion, D0), at the time of myeloid engraftment (Engraftment) and at monthly intervals after the day of infusion (1 Month, 2 Month, 3 Month, 6 Month). In the case of two timepoints overlapping, the sample was preferentially labeled as engraftment or month 1/2/3/6 (in that order). Sample processing was prioritized for post-transplantation time points due to their significance in monitoring complications. Samples from healthy stem cell transplant donors prior to stem cell mobilization were used as a healthy reference control.

#### Clinical Cohort 2

Adult recipients that had received HSCT at the Moffitt Cancer Centre were enrolled on a rolling basis from October 2010 to February 2018. Samples were collected at the following timepoints: the first day of immunosuppression taper, the day of immunosuppression stop, the day of taper failure (GVHD, during or after taper), and at two weeks, one month, three months, and six months after the end of taper.

### Engraftment definition

Neutrophil engraftment was considered when blood samples contained an absolute neutrophil count greater or equal than 500 cells per microliter blood on two separate measurements. Median day of engraftment was 19 days after transplant.

### Recipient Categorization

In cohort 1, recipients were categorized into three groups: healthy stem cell donors, early complication-free, and early complication-present. Healthy stem cell donors were individuals who provided stem cell products for recipients in this study. Complication-present recipients were HSCT recipients that were diagnosed with any of the following complications within 9 months of infusion: relapse, graft failure, acute GVHD (aGVHD), chronic GVHD (cGVHD), and BK viral relapse (BKVR). Complication-free recipients were HCT recipients that were not diagnosed with a complication within 9 months of infusion. Medical records of each recipient in the complication-free cohort were reviewed to confirm that none of these recipients experienced CMV or EBV reactivation, veno occlusive disease (VOD), hemolytic uremic syndrome (HUS) or serious infections after engraftment. If one of those complications were experienced, that recipient was categorized as early complication-present. Acute and chronic GVHD were diagnosed and graded using NIH consensus criteria (*8*, *41*, *42*).

In cohort 2, recipients were grouped into those that succeeded taper (Success) and those that failed taper (Failure). Failure was characterized as chronic GVHD or late acute GVHD occurrence either during taper or after ending taper. GVHD were diagnosed and graded using NIH consensus criteria (*8*, *41*, *42*).

### Complication Diagnosis Criteria

BK Viral Reactivation was identified using a commercial real-time PCR assay on urine and blood samples (Eurofins Viracor, LLC, Lenexa, KS). Relapse, graft failure, and GVHD: Medical records, including medical oncology notes, clinical laboratory tests and pathology reports were retrospectively reviewed. Data was captured in a clinical management system. recipients were monitored for relapse, graft failure, GVHD, infections and other complications during and after hematopoietic stem cell transplantation. Monitoring involved clinical histories, physical examinations, and regular laboratory assessments to track recipients’ progress and identify any potential complications. Supplementary biopsies were conducted to confirm diagnosis of relapse. Data collection was conducted retrospectively.

### Sample Collection

Cohort 1 (DFCI) - Blood samples were collected through standard venipuncture in ethylenediaminetetraacetic acid tubes (Becton Dickinson, reference No. 366643). Plasma was extracted through blood centrifugation (2,000 rpm for 10 min using a Beckman Coulter Allegra 6R centrifuge) and stored in 0.5- to 2-mL aliquots at −80 °C. Plasma samples were shipped from the Dana-Farber Cancer Institute to Cornell University on dry ice.

Cohort 2 (MCC) - Blood samples were collected through standard venipuncture in CPT-Na citrate tubes (Becton Dickinson, reference No. 362753). Plasma was extracted through double blood centrifugation (first: 1,500 g for 20 min at room temperature, second: 609 g for 5 min at room temperature) and stored in 1mL aliquots at −80 °C. Plasma samples were shipped from the Moffitt Cancer Center to Cornell University on dry ice.

### Sample processing and sequencing

Samples were processed using methods described previously (*27*), with the following modifications: sequencing was done on an Illumina NextSeq or NovaSeq and reads were trimmed to 61 base pairs.

### Sample quality filtering

Quality filtering was done as previously described (*27*), with the following modifications: Samples were removed from analysis if either the intron to exon ratio was greater than 3 or if the 5’-3’ read alignment ratio bias was greater than 3 z-scores from the mean.

### Correlation to healthy stem cell donors

Average Pearson correlation was calculated between each sample and each healthy stem cell donor sample using the top 500 most variable genes after variance stabilizing transformation in the Engraftment and one, two, three, and six month follow up time points. For healthy stem cell donor samples, correlation to self was excluded from the average calculation. Variance stabilizing transformation was performed using the DESeq2 v*arianceStabilizingTransformation* function. The same analysis was performed for within-group correlations at each time point and correlation to self was excluded from the average calculation.

### Distance to healthy stem cell donors

We introduce a method to measure distance of cell type of origin profiles in cfRNA that accurately reflects the unique characteristics of cell type profiles by considering two critical sources of variation: the inherent variability among different cell types and the correlation patterns between them. This approach ensures that the metric: 1) down-weights the difference for cell types with higher variability and 2) avoids overestimating differences caused by cell types that co-vary within the control population. The process involves several steps: 1) Computing the mean and covariance matrix for cell type fractions from healthy stem cell donors. Given the typical scenario where the number of cell types exceeds the sample size of controls, we apply the nearPD function from the R Matrix package to project the covariance matrix to the nearest positive-definite matrix. 2) Computing the square root of the Mahalanobis distance between the cell type fraction of the query sample and the mean of the healthy stem cell donors under the inverse of the projected covariance matrix:

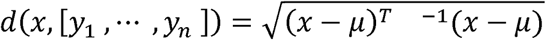

where *x* ∊ *R^k^* denotes the cell type fraction of the query sample across k cell types, *y*_1_, …, *y_n_* ∊ *R^k^* denote the cell type fraction of control samples, μ, and ∑ are the mean and projected covariance matrix of samples *y*. The implementation is based on the Mahalanobis function provided by the R matrixcalc package. The cfRNAdist R package is provided at https://github.com/Team-De-Vlaminck/cfRNAdist.git.

### Cell counts and cell type of origin deconvolution

Cell counts were obtained from blood samples collected at the same time as the plasma for cfRNA analysis. Blood samples were collected in EDTA containing tubes and subsequently analyzed at the clinical laboratories of Brigham and Women’s Hospital or DFCI (Cohort 1) or Moffitt Cancer Center (Cohort 2) using automated image-based cell counters. Automated cell counters provided WBC differential counts as well as complete blood cell counts.

Deconvolution of cfRNA to obtain cell type fractions was done using BayesPrism(v3.0) (*22*) using Tabula Sapiens as a single cell reference (*23*). For lymphocytes, the sum of deconvolution results for B cells, T cells, plasma cells, NK cells, and innate lymphoid cells was used. For leukocytes, the sum of deconvolution results for all lymphocytes, along with neutrophils, basophils, mast cells, macrophages, mature conventional dendritic cells, and monocytes was used.

### Differential cell type of origin and transcript abundance analysis

Differentially abundant RNAs were calculated using DESeq2 (*43*). Prior to analysis, transcripts with a baseMean < 5 were removed, and p-values were adjusted using the Benjamini-Hochberg method. Differential cell type of origin analysis was performed using unpaired Wilcoxon tests and p-values were adjusted using the Benjamini-Hochberg method. Pathway analysis was performed using Qiagen Ingenuity Pathway Analysis (IPA).

Longitudinal differential abundance analysis was performed using DESeq2 (*43*) and p-values were adjusted using the Benjamini-Hochberg methods. Sampling time point and day relative to aGVHD diagnosis were used as covariates.

### Machine learning modeling

For our machine learning classification models, we first normalized the counts using the trimmed means of the M-values approach (TMM) normalized to counts per million (CPM). We then conducted 101 repeated stratified train-test splits without replacement to train and evaluate LASSO regression models, assigning 80% of the samples to training and 20% to testing in each split. For the immunosuppression taper analysis, we used stratified group splitting to ensure that all samples from the same recipient were placed in either the training or testing set, thereby preventing data leakage.

### Single cell analysis

The Tabula Sapiens scRNA-seq atlas preprocessed h5ad file containing cell labels and normalized counts was downloaded from CellxGene and analyzed in Python using the Scanpy package. Only cells using the 10x genomics 3’ v3 technology were analyzed. The Scanpy pl.umap function was used for visualization, and cells labelled as “endothelial cell of lymphatic vessel” or “erythroid progenitor cell” were plotted with increased size for visualization purposes. Pre-normalized counts were utilized to show expression.

### Statistical analyses

The programming language R (v4.1.0) was utilized for all statistical analyses. Statistical significance was assessed through two-sided Wilcoxon signed-rank tests and Mann-Whitney U tests, unless specified otherwise. Machine learning algorithms were trained using the Caret R package and pipelines were run using the Snakemake workflow management system. In boxplots, boxes denote the 25th and 75th percentiles, the band within the box signifies the median, and whiskers extend to 1.5 times the interquartile range of the hinge. The alignment of all sequencing data was performed against the GRCh38 Gencode v38 Primary Assembly, with feature counting conducted using the GRCh38 Gencode v38 Primary Assembly Annotation.

## Data Availability

De-identified RNA-seq count matrices have been uploaded to the NCBI (National Center for Biotechnology Information) GEO (Gene Expression Omnibus) database and will be publicly available upon publication.

## Acknowledgments

This work would not have been possible without the vision, guidance, and leadership of the late Dr. Francisco M. Marty, who was instrumental in launching this study. The co-authors are grateful for his friendship, mentorship, and for everything he taught us. We thank the patients and their families for their help to further our understanding hematopoietic stem cell transplantation.

## Author contributions

Conceived and designed the study: CJL, MPC, JR, JP, and IDV

Performed sequencing experiments: AB, JL, and EB

Identified and collected recipient samples and clinical metadata: IHGB, MPC, JP, and JR

Provided input for data analysis: MPC, TC, DEL

Analyzed sequencing data: CJL, GA, TC, IDV

Supervised the study: MPC, JP, JR, IDV

Wrote the manuscript and prepared the figures: CJL, IDV

All authors read and edited the manuscript and agreed to its contents.

## Data and materials availability

De-identified RNA-seq count matrices have been uploaded to the NCBI (National Center for Biotechnology Information) GEO (Gene Expression Omnibus) database and will be publicly available upon publication (GSE268164). All code has been deposited on GitHub (https://github.com/conorloy/cfrnaHSCT) and will be available upon publication.

## Supplementary Materials

**Fig. S1.**
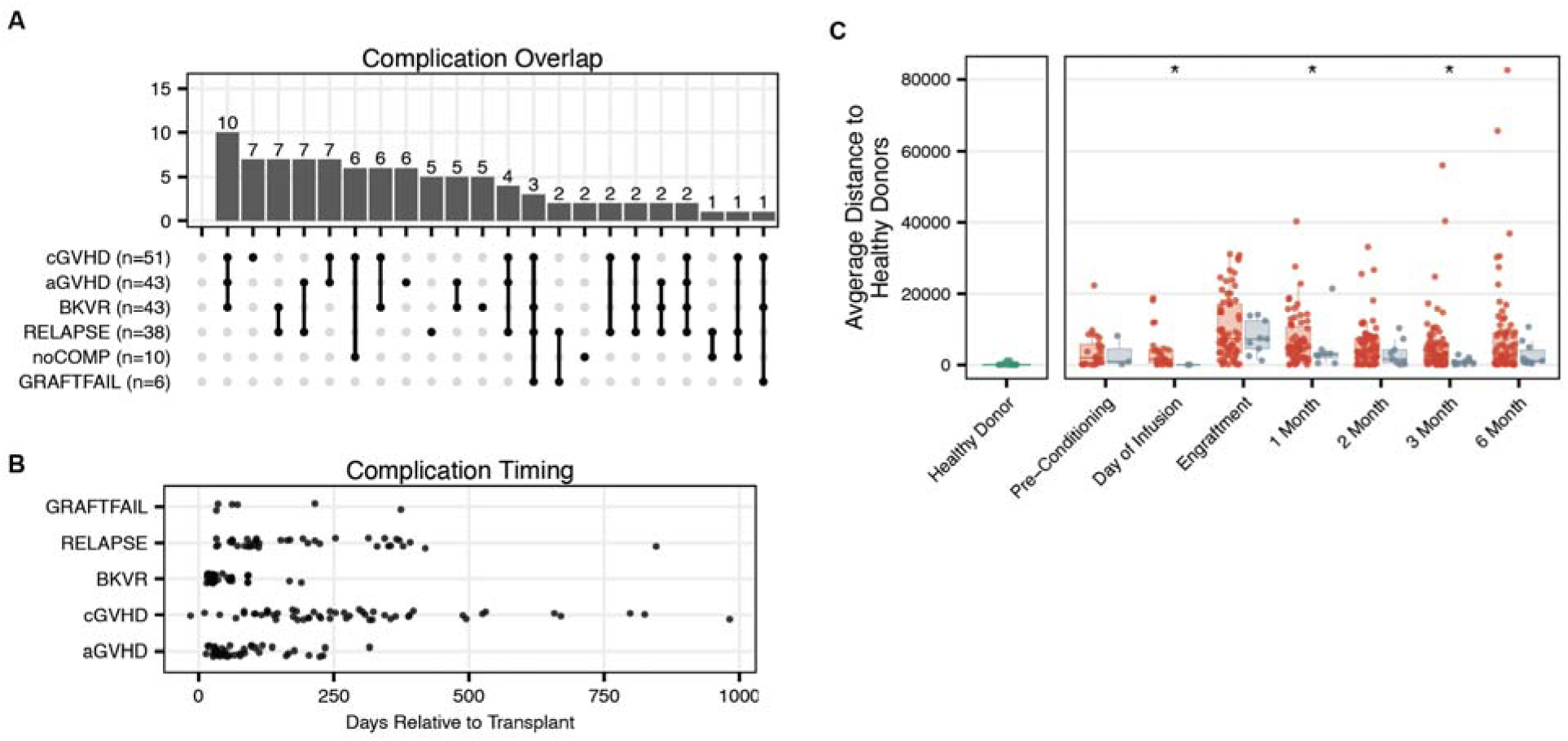
(**A**) Upset plot of complications. (**B**) Timing of complications relative to transplantation. (**C**) Distance of cfRNA cell type of origin profiles to healthy stem cell donors of samples from with and without complications. Asterisks indicate statistical significance: ns, non-significant; *, p < 0.1; **, p < 0.01; ***, p < 0.001; ****, p < 0.001.

**Fig. S2.**
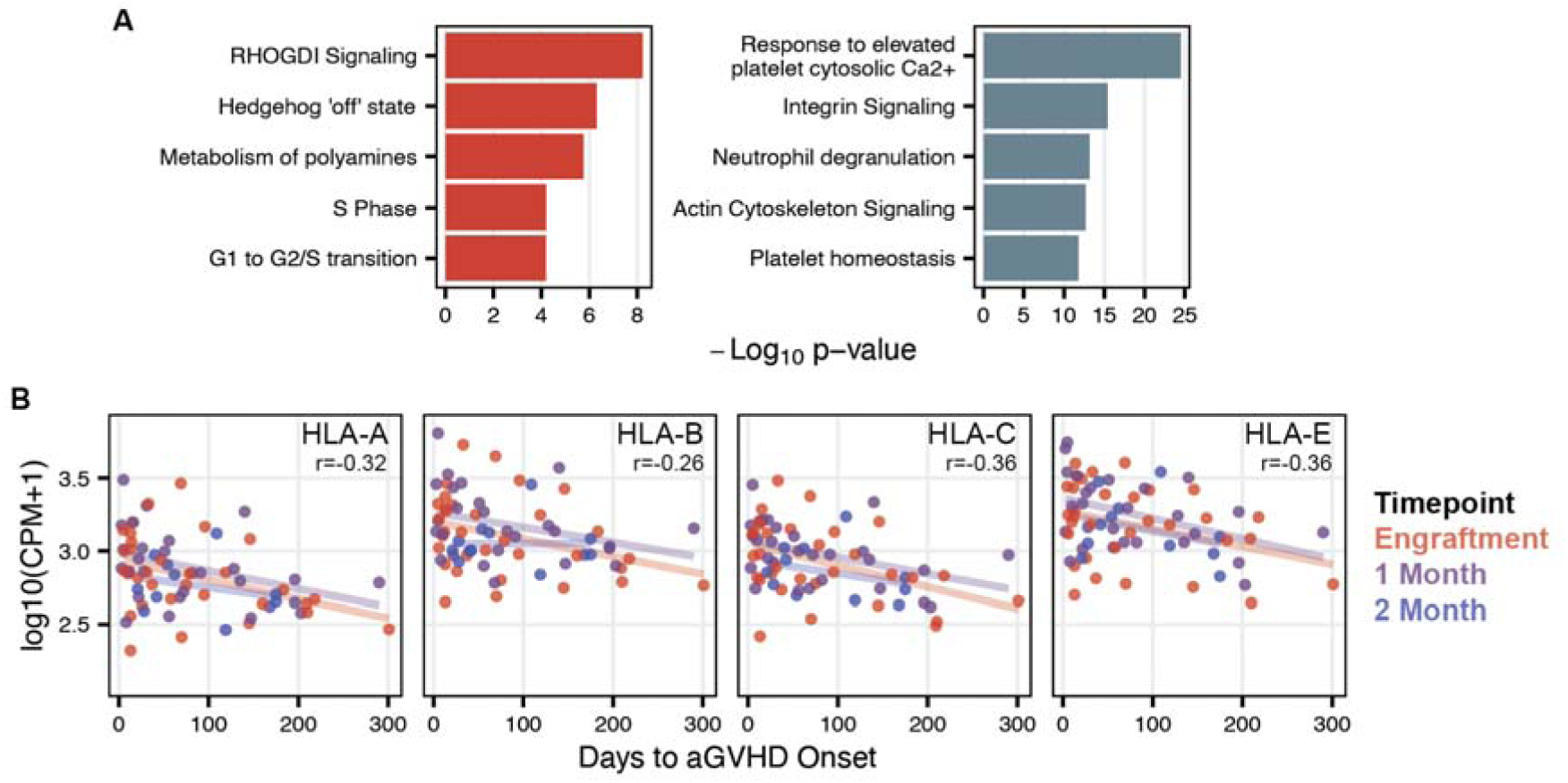
(**A**) Top five significantly enriched pathways in the set of genes found to be differentially abundant between engraftment samples from patients who do and do not develop aGVHD (Qiagen, IPA). (**B**) Major Histocompatibility Complex, Class I - A, B, C and E (*HLA-A*, *HLA-B*, *HLA-C*, and *HLA-E*) transcript counts per million relative to days to aGVHD diagnosis in samples at the engraftment, one-month, and two-month time points. Pearson correlation calculated for all samples. Best fit line plotted for each timepoint.

**Table S1.** Abbreviations: M: male; F: female; AA: aplastic anemia; AML: acute myeloid leukemia; ALL: acute lymphoid leukemia; CLL: chronic lymphocytic leukemia; CMML: chronic myelomonocytic leukemia; MDS: myelodysplastic syndrome; MM: multiple myeloma; MPS: myelodysplastic syndrome; PBSC: peripheral blood stem cell; HLA: human leukocyte antigen; Tac: tacrolimus; Sir: sirolimus/ rapamycin; MTX: methotrexate; MMF: mycophenolate mofetil

|  |  |  |  |
| --- | --- | --- | --- |
|  | N |  | N |
| **Total** | 96 |  | 96 |
| **Age, median (range)** | 63 (19, 78) | **Acute GVHD prophylaxis** |  |
| **recipient-Donor Gender** |  | Tac-Sir | 5 |
| M-M | 41 | MMF-Tac | 41 |
| M-F | 16 | MTX-Tac | 28 |
| F-M | 16 | MTX-Tac-Sir | 22 |
| F-F | 23 | **Stem Cell Source** |  |
| **Diagnosis** |  | PBSC | 73 |
| AA | 4 | Bone marrow | 19 |
| ALL | 11 | Umbilical cord | 4 |
| AML | 35 | **Acute GvHD grade** |  |
| Acute Leukemia | 2 | 0 | 55 |
| CLL | 1 | 1 | 20 |
| CMML | 3 | 2 | 13 |
| Lymphoma | 9 | 3 | 3 |
| MDS | 17 | 4 | 5 |
| MM | 1 | **Chronic GvHD severity** |  |
| MPS | 5 | None | 44 |
| Other | 8 | Mild | 22 |
| **Donor HLA Type** |  | Moderate | 25 |
| Matched related | 29 | Severe | 5 |
| Matched unrelated | 47 |  |  |
| Mismatched unrelated | 20 |  |  |

**Table S2.** Acute GvHD grade and Chronic GvHD severity recorded from after the start of immunosuppression taper. Abbreviations: M: male; F: female; ALL: acute lymphoid leukemia; AML: acute myeloid leukemia; BPDCN: Blastic Plasmacytoid Dendritic Cell Neoplasm CLL: chronic lymphocytic leukemia; CMML: chronic myelomonocytic leukemia; MDS: myelodysplastic syndrome; MM: multiple myeloma; MPD: myeloproliferative disorder; NHL: Non-Hodgkin Lymphoma; SAA: Severe aplastic anemia; HLA: human leukocyte antigen; Tac: tacrolimus; Sir: sirolimus/ rapamycin; MTX: methotrexate; MMF: mycophenolate mofetil, PTCy: Post-transplantation cyclophosphamide, PBSC: peripheral blood stem cell;

|  | N |  | N |
| --- | --- | --- | --- |
| **Total** | 54 |  | 54 |
| **Age, median (range)** | 60 (32, 72) | **Acute GVHD prophylaxis** |  |
| **recipient-Donor Gender** |  | Tac-Sir | 35 |
| M-M | 15 | MMF-Tac | 3 |
| M-F | 14 | MTX-Tac | 13 |
| F-M | 13 | MMF-Tac-PTCy | 1 |
| F-F | 11 | Sir-MMF-PTCy | 1 |
| **Diagnosis** |  | **Stem Cell Source** |  |
| ALL | 4 | PBSC | 152 |
| AML | 18 | Bone marrow | 1 |
| BPDCN | 1 | **Acute GvHD grade** |  |
| CLL | 2 | 0 | 51 |
| CMML | 4 | 1 | 1 |
| MDS | 10 | 2 | 1 |
| MM | 7 | 3 | 1 |
| MPD | 2 | 4 | 0 |
| NHL | 3 | **Chronic GvHD severity** |  |
| SAA | 2 | None | 31 |
| Other | 0 | Mild | 6 |
| **Donor HLA Type** |  | Moderate | 14 |
| Match | 50 | Severe | 3 |
| Mismatched | 3 |  |  |

**Data S1. (separate file)**

DESeq2 output from aGVHD+ vs aGVHD- using pre-diagnosis engraftment timepoint samples.

**Data S2. (separate file)**

DESeq2 output analyzing time to aGVHD diagnosis from sample collection.

**Data S3. (separate file)**

DESeq2 output from cGVHD/L-aGVHD positive vs negative patients using three- and six-month time point samples.

**Data S4. (separate file)**

DESeq2 output from taper success vs taper failure.

## References and Notes

1. N. Cieri, K. Maurer, C. J. Wu, 60 Years Young: The Evolving Role of Allogeneic Hematopoietic Stem Cell Transplantation in Cancer Immunotherapy. Cancer Research 81, 4373–4384 (2021).

2. P. L. Weiden, N. Flournoy, E. D. Thomas, R. Prentice, A. Fefer, C. D. Buckner, R. Storb, Antileukemic Effect of Graft-versus-Host Disease in Human Recipients of Allogeneic-Marrow Grafts. N Engl J Med 300, 1068–1073 (1979).

3. M. M. Horowitz, R. P. Gale, P. M. Sondel, J. M. Goldman, J. Kersey, H. J. Kolb, A. A. Rimm, O. Ringdén, C. Rozman, B. Speck, Graft-versus-leukemia reactions after bone marrow transplantation. Blood 75, 555–562 (1990).

4. C. Annaloro, F. Serpenti, G. Saporiti, G. Galassi, F. Cavallaro, F. Grifoni, M. Goldaniga, L. Baldini, F. Onida, Viral Infections in HSCT: Detection, Monitoring, Clinical Management, and Immunologic Implications. Front. Immunol. 11, 569381 (2021).

5. F. Kreidieh, I. Abou Dalle, N. Moukalled, J. El-Cheikh, E. Brissot, M. Mohty, A. Bazarbachi, Relapse after allogeneic hematopoietic stem cell transplantation in acute myeloid leukemia: an overview of prevention and treatment. Int J Hematol 116, 330–340 (2022).

6. C. McDonald-Hyman, L. A. Turka, B. R. Blazar, Advances and challenges in immunotherapy for solid organ and hematopoietic stem cell transplantation. Sci. Transl. Med. 7 (2015).

7. R. Zeiser, B. R. Blazar, Acute Graft-versus-Host Disease - Biologic Process, Prevention, and Therapy. N Engl J Med 377, 2167–2179 (2017).

8. M. H. Jagasia, H. T. Greinix, M. Arora, K. M. Williams, D. Wolff, E. W. Cowen, J. Palmer, D. Weisdorf, N. S. Treister, G.-S. Cheng, H. Kerr, P. Stratton, R. F. Duarte, G. B. McDonald, Y. Inamoto, A. Vigorito, S. Arai, M. B. Datiles, D. Jacobsohn, T. Heller, C. L. Kitko, S. A. Mitchell, P. J. Martin, H. Shulman, R. S. Wu, C. S. Cutler, G. B. Vogelsang, S. J. Lee, S. Z. Pavletic, M. E. D. Flowers, National Institutes of Health Consensus Development Project on Criteria for Clinical Trials in Chronic Graft-versus-Host Disease: I. The 2014 Diagnosis and Staging Working Group Report. Biology of Blood and Marrow Transplantation 21, 389-401.e1 (2015).

9. J. A. Pidala, T. A. Gooley, L. Luznik, B. R. Blazar, Chronic graft-versus-host disease: unresolved complication or ancient history? Blood 144, 1363–1373 (2024).

10. G. Socié, L. S. Kean, R. Zeiser, B. R. Blazar, Insights from integrating clinical and preclinical studies advance understanding of graft-versus-host disease. Journal of Clinical Investigation 131, e149296 (2021).

11. T. Pandey, S. Maximin, P. Bhargava, Imaging of complications from hematopoietic stem cell transplant. Indian Journal of Radiology and Imaging 24, 327–338 (2014).

12. S. Jodele, S. M. Davies, A. Lane, J. Khoury, C. Dandoy, J. Goebel, K. Myers, M. Grimley, J. Bleesing, J. El-Bietar, G. Wallace, R. S. Chima, Z. Paff, B. L. Laskin, Diagnostic and risk criteria for HSCT-associated thrombotic microangiopathy: a study in children and young adults. Blood 124, 645–653 (2014).

13. J. L. M. Ferrara, M. S. Chaudhry, GVHD: biology matters. Blood Adv 2, 3411–3417 (2018).

14. S. Paczesny, Discovery and validation of graft-versus-host disease biomarkers. Blood 121, 585–594 (2013).

15. T. Toubai, J. Magenau, Immunopathology and biology-based treatment of steroid-refractory graft-versus-host disease. Blood 136, 429–440 (2020).

16. C. Loy, L. Ahmann, I. De Vlaminck, W. Gu, Liquid Biopsy Based on Cell-Free DNA and RNA. Annu. Rev. Biomed. Eng. 26, annurev-bioeng-110222-111259 (2024).

17. A. P. Cheng, M. P. Cheng, C. J. Loy, J. S. Lenz, K. Chen, S. Smalling, P. Burnham, K. M. Timblin, J. L. Orejas, E. Silverman, P. Polak, F. M. Marty, J. Ritz, I. De Vlaminck, Cell-free DNA profiling informs all major complications of hematopoietic cell transplantation. Proceedings of the National Academy of Sciences 119, e2113476118 (2022).

18. B. Avni, D. Neiman, E. Shaked, O. Gal-Rosenberg, S. Grisariu, M. Kuzli, I. Avni, A. Fracchia, P. Stepensky, T. Zuckerman, A. Lev-Sagie, I. Fox-Fisher, S. Piyanzin, J. Moss, S. J. Salpeter, B. Glaser, R. Shemer, Y. Dor, Chronic graft-versus-host disease detected by tissue-specific cell-free DNA methylation biomarkers. Journal of Clinical Investigation 134, e163541 (2024).

19. Y. Pang, T. E. Andargie, M. K. Jang, H. Kong, W. Park, T. Hill, N. Redekar, Y.-P. Fu, D. A. Parth, N. G. Holtzman, S. Z. Pavletic, S. Agbor-Enoh, Chronic graft-versus-host disease is characterized by high levels and distinctive tissue-of-origin patterns of cell-free DNA. iScience 26, 108160 (2023).

20. A. Ibarra, J. Zhuang, Y. Zhao, N. S. Salathia, V. Huang, A. D. Acosta, J. Aballi, S. Toden, A. P. Karns, I. Purnajo, J. R. Parks, L. Guo, J. Mason, D. Sigal, T. S. Nova, S. R. Quake, M. Nerenberg, Non-invasive characterization of human bone marrow stimulation and reconstitution by cell-free messenger RNA sequencing. Nat Commun 11, 400 (2020).

21. W. W. B. Goh, L. Wong, Dealing with Confounders in Omics Analysis. Trends in Biotechnology 36, 488–498 (2018).

22. T. Chu, Z. Wang, D. Pe’er, C. G. Danko, Cell type and gene expression deconvolution with BayesPrism enables Bayesian integrative analysis across bulk and single-cell RNA sequencing in oncology. Nat Cancer 3, 505–517 (2022).

23. Tabula Sapiens Consortium*, R. C. Jones, J. Karkanias, M. A. Krasnow, A. O. Pisco, S. R. Quake, J. Salzman, N. Yosef, B. Bulthaup, P. Brown, W. Harper, M. Hemenez, R. Ponnusamy, A. Salehi, B. A. Sanagavarapu, E. Spallino, K. A. Aaron, W. Concepcion, J. M. Gardner, B. Kelly, N. Neidlinger, Z. Wang, S. Crasta, S. Kolluru, M. Morri, A. O. Pisco, S. Y. Tan, K. J. Travaglini, C. Xu, M. Alcántara-Hernández, N. Almanzar, J. Antony, B. Beyersdorf, D. Burhan, K. Calcuttawala, M. M. Carter, C. K. F. Chan, C. A. Chang, S. Chang, A. Colville, S. Crasta, R. N. Culver, I. Cvijović, G. D’Amato, C. Ezran, F. X. Galdos, A. Gillich, W. R. Goodyer, Y. Hang, A. Hayashi, S. Houshdaran, X. Huang, J. C. Irwin, S. Jang, J. V. Juanico, A. M. Kershner, S. Kim, B. Kiss, S. Kolluru, W. Kong, M. E. Kumar, A. H. Kuo, R. Leylek, B. Li, G. B. Loeb, W.-J. Lu, S. Mantri, M. Markovic, P. L. McAlpine, A. de Morree, M. Morri, K. Mrouj, S. Mukherjee, T. Muser, P. Neuhöfer, T. D. Nguyen, K. Perez, R. Phansalkar, A. O. Pisco, N. Puluca, Z. Qi, P. Rao, H. Raquer-McKay, N. Schaum, B. Scott, B. Seddighzadeh, J. Segal, S. Sen, S. Sikandar, S. P. Spencer, L. C. Steffes, V. R. Subramaniam, A. Swarup, M. Swift, K. J. Travaglini, W. Van Treuren, E. Trimm, S. Veizades, S. Vijayakumar, K. C. Vo, S. K. Vorperian, W. Wang, H. N. W. Weinstein, J. Winkler, T. T. H. Wu, J. Xie, A. R. Yung, Y. Zhang, A. M. Detweiler, H. Mekonen, N. F. Neff, R. V. Sit, M. Tan, J. Yan, G. R. Bean, V. Charu, E. Forgó, B. A. Martin, M. G. Ozawa, O. Silva, S. Y. Tan, A. Toland, V. N. P. Vemuri, S. Afik, K. Awayan, O. B. Botvinnik, A. Byrne, M. Chen, R. Dehghannasiri, A. M. Detweiler, A. Gayoso, A. A. Granados, Q. Li, G. Mahmoudabadi, A. McGeever, A. de Morree, J. E. Olivieri, M. Park, A. O. Pisco, N. Ravikumar, J. Salzman, G. Stanley, M. Swift, M. Tan, W. Tan, A. J. Tarashansky, R. Vanheusden, S. K. Vorperian, P. Wang, S. Wang, G. Xing, C. Xu, N. Yosef, M. Alcántara-Hernández, J. Antony, C. K. F. Chan, C. A. Chang, A. Colville, S. Crasta, R. Culver, L. Dethlefsen, C. Ezran, A. Gillich, Y. Hang, P.-Y. Ho, J. C. Irwin, S. Jang, A. M. Kershner, W. Kong, M. E. Kumar, A. H. Kuo, R. Leylek, S. Liu, G. B. Loeb, W.-J. Lu, J. S. Maltzman, R. J. Metzger, A. de Morree, P. Neuhöfer, K. Perez, R. Phansalkar, Z. Qi, P. Rao, H. Raquer-McKay, K. Sasagawa, B. Scott, R. Sinha, H. Song, S. P. Spencer, A. Swarup, M. Swift, K. J. Travaglini, E. Trimm, S. Veizades, S. Vijayakumar, B. Wang, W. Wang, J. Winkler, J. Xie, A. R. Yung, S. E. Artandi, P. A. Beachy, M. F. Clarke, L. C. Giudice, F. W. Huang, K. C. Huang, J. Idoyaga, S. K. Kim, M. Krasnow, C. S. Kuo, P. Nguyen, S. R. Quake, T. A. Rando, K. Red-Horse, J. Reiter, D. A. Relman, J. L. Sonnenburg, B. Wang, A. Wu, S. M. Wu, T. Wyss-Coray, The Tabula Sapiens: A multiple-organ, single-cell transcriptomic atlas of humans. Science 376, eabl4896 (2022).

24. J. Pidala, M. Martens, C. Anasetti, J. Carreras, M. Horowitz, S. J. Lee, J. Antin, C. Cutler, B. Logan, Factors Associated With Successful Discontinuation of Immune Suppression After Allogeneic Hematopoietic Cell Transplantation. JAMA Oncol 6, e192974 (2020).

25. G. L. Chen, L. Onstad, P. J. Martin, P. Carpenter, J. Pidala, S. Arai, C. Cutler, B. K. Hamilton, S. J. Lee, M. Arora, Durable discontinuation of systemic therapy in patients affected by chronic graft-versus-host disease. Haematologica 108, 483–489 (2023).

26. A. Chang, C. J. Loy, J. S. Lenz, A. Steadman, A. Andama, N. V. Nhung, C. Yu, W. Worodria, C. M. Denkinger, P. Nahid, A. Cattamanchi, I. De Vlaminck, Circulating Cell-Free RNA in Blood as a Host Response Biomarker for the Detection of Tuberculosis. medRxiv, 2023.01.11.23284433 (2023).

27. C. J. Loy, A. Sotomayor-Gonzalez, V. Servellita, J. Nguyen, J. Lenz, S. Bhattacharya, M. E. Williams, A. P. Cheng, A. Bliss, P. Saldhi, N. Brazer, J. Streithorst, W. Suslovic, C. J. Hsieh, B. Bahar, N. Wood, A. Foresythe, A. Gliwa, K. Bhakta, M. A. Perez, L. Hussaini, E. J. Anderson, A. Chahroudi, M. Delaney, A. J. Butte, R. L. DeBiasi, C. A. Rostad, I. De Vlaminck, C. Y. Chiu, Nucleic acid biomarkers of immune response and cell and tissue damage in children with COVID-19 and MIS-C. Cell Reports Medicine, 101034 (2023).

28. Y. Wang, J. Li, L. Zhang, H.-X. Sun, Z. Zhang, J. Xu, Y. Xu, Y. Lin, A. Zhu, Y. Luo, H. Zhou, Y. Wu, S. Lin, Y. Sun, F. Xiao, R. Chen, L. Wen, W. Chen, F. Li, R. Ou, Y. Zhang, T. Kuo, Y. Li, L. Li, J. Sun, K. Sun, Z. Zhuang, H. Lu, Z. Chen, G. Mai, J. Zhuo, P. Qian, J. Chen, H. Yang, J. Wang, X. Xu, N. Zhong, J. Zhao, J. Li, J. Zhao, X. Jin, Plasma cell-free RNA characteristics in COVID-19 patients. Genome Res 32, 228–241 (2022).

29. K. L. Kalantar, L. Neyton, M. Abdelghany, E. Mick, A. Jauregui, S. Caldera, P. H. Serpa, R. Ghale, J. Albright, A. Sarma, A. Tsitsiklis, A. Leligdowicz, S. A. Christenson, K. Liu, K. N. Kangelaris, C. Hendrickson, P. Sinha, A. Gomez, N. Neff, A. Pisco, S. B. Doernberg, J. L. Derisi, M. A. Matthay, C. S. Calfee, C. R. Langelier, Integrated host-microbe plasma metagenomics for sepsis diagnosis in a prospective cohort of critically ill adults. Nat Microbiol 7, 1805–1816 (2022).

30. N. Chalasani, S. Toden, J. J. Sninsky, R. P. Rava, J. V. Braun, S. Gawrieh, J. Zhuang, M. Nerenberg, S. R. Quake, T. Maddala, Noninvasive stratification of nonalcoholic fatty liver disease by whole transcriptome cell-free mRNA characterization. Am J Physiol Gastrointest Liver Physiol 320, G439–G449 (2021).

31. K. R. Schultz, A. Kariminia, B. Ng, S. Abdossamadi, M. Lauener, E. R. Nemecek, J. T. Wahlstrom, C. L. Kitko, V. A. Lewis, T. Schechter, D. A. Jacobsohn, A. C. Harris, M. A. Pulsipher, H. Bittencourt, S. W. Choi, E. H. Caywood, K. A. Kasow, M. Bhatia, B. R. Oshrine, A. Flower, S. Chaudhury, D. Coulter, J. H. Chewning, M. Joyce, S. Savasan, A. B. Pawlowska, G. C. Megason, D. Mitchell, A. C. Cheerva, A. Lawitschka, S. Azadpour, E. Ostroumov, P. Subrt, A. Halevy, S. Mostafavi, G. D. E. Cuvelier, Immune profile differences between chronic GVHD and late acute GVHD: results of the ABLE/PBMTC 1202 studies. Blood 135, 1287–1298 (2020).

32. A. Kariminia, S. G. Holtan, S. Ivison, J. Rozmus, M.-J. Hebert, P. J. Martin, S. J. Lee, D. Wolff, P. Subrt, S. Abdossamadi, S. Sung, J. Storek, M. Levings, M. Aljurf, M. Arora, C. Cutler, G. Gallagher, J. Kuruvilla, J. Lipton, T. J. Nevill, L. F. Newell, T. Panzarella, J. Pidala, G. Popradi, D. Szwajcer, J. Tay, C. L. Toze, I. Walker, S. Couban, B. E. Storer, K. R. Schultz, Heterogeneity of chronic graft-versus-host disease biomarkers: association with CXCL10 and CXCR3+ NK cells. Blood 127, 3082–3091 (2016).

33. S. Ghimire, D. Weber, E. Mavin, X. N. Wang, A. M. Dickinson, E. Holler, Pathophysiology of GvHD and Other HSCT-Related Major Complications. Front. Immunol. 8 (2017).

34. C. M. Westhoff, The Structure and Function of the Rh Antigen Complex. Seminars in Hematology 44, 42–50 (2007).

35. M. Yazer, D. Triulzi, J. Sperry, A. Corcos, J. Seheult, Rate of RhD-alloimmunization after the transfusion of RhD-positive red blood cell containing products among injured patients of childbearing age: single center experience and narrative literature review. Hematology 26, 321–327 (2021).

36. J. Cid, M. Lozano, Matching for the D antigen in haematopoietic progenitor cell transplantation: definition and clinical outcomes. Blood Transfusion, doi: 10.2450/2014.0238-13 (2014).

37. T. Konuma, N. Uchida, W. Takeda, N. Doki, S. Yoshihara, T. Nishida, T. Kuriyama, M. Tanaka, H. Ohigashi, H. Nakamae, Y. Katayama, S. Ota, Y. Hashii, F. Ishimaru, T. Fukuda, M. Ohbiki, Y. Atsuta, RhD mismatch does not affect haematopoietic recovery, graft versus host disease and survival in allogeneic haematopoietic cell transplantation: A Japanese registry based study. Vox Sanguinis 119, 612–618 (2024).

38. A. Mijovic, Alloimmunization to RhD antigen in RhD-incompatible haemopoietic cell transplants with non-myeloablative conditioning. Vox Sang 83, 358–362 (2002).

39. I. Comerford, Y. Harata-Lee, M. D. Bunting, C. Gregor, E. E. Kara, S. R. McColl, A myriad of functions and complex regulation of the CCR7/CCL19/CCL21 chemokine axis in the adaptive immune system. Cytokine Growth Factor Rev 24, 269–283 (2013).

40. S. Changsirikulchai, D. Myerson, K. A. Guthrie, G. B. McDonald, C. E. Alpers, S. R. Hingorani, Renal Thrombotic Microangiopathy after Hematopoietic Cell Transplant: Role of GVHD in Pathogenesis. Clinical Journal of the American Society of Nephrology 4, 345–353 (2009).

41. on behalf of the EBMT (European Society for Blood and Marrow Transplantation) Transplant Complications Working Party and the “EBMT−NIH (National Institutes of Health)−CIBMTR (Center for International Blood and Marrow Transplant Research) GvHD Task Force,” H. M. Schoemans, S. J. Lee, J. L. Ferrara, D. Wolff, J. E. Levine, K. R. Schultz, B. E. Shaw, M. E. Flowers, T. Ruutu, H. Greinix, E. Holler, G. Basak, R. F. Duarte, S. Z. Pavletic, EBMT−NIH−CIBMTR Task Force position statement on standardized terminology & guidance for graft-versus-host disease assessment. Bone Marrow Transplant 53, 1401–1415 (2018).

42. A. C. Harris, R. Young, S. Devine, W. J. Hogan, F. Ayuk, U. Bunworasate, C. Chanswangphuwana, Y. A. Efebera, E. Holler, M. Litzow, R. Ordemann, M. Qayed, A. S. Renteria, R. Reshef, M. Wölfl, Y.-B. Chen, S. Goldstein, M. Jagasia, F. Locatelli, S. Mielke, D. Porter, T. Schechter, Z. Shekhovtsova, J. L. M. Ferrara, J. E. Levine, International, Multicenter Standardization of Acute Graft-versus-Host Disease Clinical Data Collection: A Report from the Mount Sinai Acute GVHD International Consortium. Biol Blood Marrow Transplant 22, 4–10 (2016).

43. M. I. Love, W. Huber, S. Anders, Moderated estimation of fold change and dispersion for RNA-seq data with DESeq2. Genome Biol 15, 550 (2014).

